# Facial Palsy and the eyes: Epidemiology, Tear Film, and Meibomian gland dysfunction; a comparative analysis

**DOI:** 10.1101/2023.07.24.23293013

**Authors:** Caroline Guerrero-de Ferran, Jocelyn I. Rivera-Alvarado, Daniel Bastán-Fabián, Alfredo Del Castillo-Morales, Fabiola Velázquez-Valenzuela, Jorge E. Valdez García

**Affiliations:** Tecnologico de Monterrey School of Medicine

**Author notes:** **Corresponding author** Jorge E. Valdez-Garcia, School of Medicine and Health Sciences, Av. Ignacio Morones Prieto 3000. Colonia Los Doctores. CP 64710., Monterrey, Nuevo Leon.

**Keywords:** Meibomian gland dysfunction, facial palsy, meibography, dry eye disease, orbicularis oculi

## Abstract

**Purpose:** To describe the correlation between facial palsy of any etiology and Meibomian gland dysfunction and to report the tear film parameters found in patients with facial palsy diagnosis.

**Design:** Observational, longitudinal, and comparative study.

**Methods:** A sample of 20 patients with unilateral facial palsy was obtained its severity was staged using the House-Brackmann scale. A dry eye evaluation using an Oculus 5M keratograph was performed, which included infrared meibography, tear breakup time, and a dry eye questionnaire. Meibographies were analyzed using ImageJ software to determine the affected area.

**Results:** 11 patients (55%) were female, mean age of diagnosis was 57.80 ± 18.28 (range of 22-80), 10 (50%) cases were due to Bell’s palsy. Tear breakup time was markedly reduced in the affected side but was statistically insignificant (p=0.2167) and tear meniscus height was much greater in the non-affected side (p=0.0199). Finally, Meibomian gland alterations were more evident in the affected side, with upper Meibomian glands having a loss of 29.55 ± 13.31 percent (p=0.0374) and lower glands presenting a loss 44.44 ± 16.9 percent (p=<0.001).

**Conclusions:** A clear difference in Meibomian gland and tear film dynamics can be observed in paretic vs non paretic sides in the same patients. Multiple mechanisms contribute to this ailment, such as an incomplete blinking pattern and Meibomian gland dysfunction due to orbicularis oculi weakness. Software based analysis also showed a greater glandular area loss when compared to clinician’s analysis, which may help aide in therapeutic decisions for patients with meibomian gland dysfunction.

## Introduction

Facial palsy (FP) is the most common cranial neuropathy in clinical practice, with an annual incidence of 30-40 cases per 100,000 people.^1-3^ If left untreated, it can result in long term physical and psychological sequelae with a marked decrease in quality of life.^4^ Diagnosis is eminently clinical by means of the House-Brackmann scale, which consists of 6 stages of increasing severity; the Sunnybrook scale is also employed and is considered the leading scale due to its more comprehensive grading scheme.^4^ There are many etiologies, such as idiopathic (Bell’s palsy), congenital, traumatic, infectious, and neoplastic.^4-6^

Dry eye disease (DED) affects approximately 100 million people worldwide but there is a massive number of undiagnosed patients.^7^ DED and Meibomian gland (MG) dysfunction (MGD) are current areas of interest in ophthalmology with an upwards trend in research. In 2007, the Tear Film and Ocular Surface Society (TFOS) presented an initiative called the Dry Eye Workshop (TFOS: DEWS).^8^ They defined DED as a multifactorial disease, characterized by the loss of tear film (TF) homeostasis manifested by ocular surface symptoms, where TF instability and hyperosmolarity, ocular surface inflammation and damage, as well as neurosensory abnormalities play a fundamental role.^9^ MGD is considered the most important cause of DED.^10,11^

Regarding the eye and orbit, FP can cause upper and lower eyelid dysfunction due to orbicularis oculi impairment, resulting in an inability to completely close the eyelids and an increase in ocular surface damage due to an exposure to air, foreign bodies, and pathogens. Likewise, TF dynamics are altered because of an uneven distribution of tears in the ocular surface and their drainage through the lacrimal punctae, increased evaporation due to an increased width of the palpebral fissure, and MGD.^3,12^ MGD due to FP is an interesting phenomenon, in which the lipid layer of the tear film is diminished due to Meibomian gland stasis. Studies have shown that these glands are surrounded by Riolan’s muscle, a subdivision of striated muscle, separate from the pretarsal orbicularis oculi muscle. ^13,14^ This muscle aids in glandular drainage and its weakness ultimately leads to obstruction and atrophy of the Meibomian glands.^2,14-16^

The importance of MGD in DED lies in the multi-layer nature of the TF. Meibomian glands secrete the lipid component of the tears, which contribute to increased lubrication, surface wettability, and tear breakup time (TBUT). ^2,8,10,16-23^ This lipid layer also helps in tear distribution in an evenly manner, as well as preventing tear spillage from the palpebral margins and sealing the eyelids during sleep.^8,15^ MGD is diagnosed based on a myriad of clinical features, such as conjunctival blood vessel ingurgitation, foamy tear film, Meibomian gland ostial dilation and increased viscosity of secretions, and corneal epithelial damage can be appreciated by slit-lamp examination. ^14,21,24,25^ Studies such as TBUT, corneal staining, Schirmer’s test, and meibography (either contact or non-contact) are of great assistance during consultation. ^6,14,26-31^

A novel tool in DED diagnosis is the JENVIS Dry Eye Report, which consists of a battery of tests such as tear meniscus height (TMH), TBUT, Dry Eye Questionnaire (DEQ), Infrared non-contact meibography, conjunctivochalasis, and conjunctival hyperemia measured by an Oculus 5M Keratograph (®K5 M; Oculus GmbH, Wetzlar, Germany). ^11-15^

## Methods

This study was approved by the local ethics committee in February 2020 according to the Helsinki declaration under the registration number: P000302-CDLPPFU. Informed consent was obtained from all participants. An observational, longitudinal, and comparative study was performed. 20 patients with a FP diagnosis were selected, all of which were referred to our institution from two metropolitan third level healthcare centers. Inclusion criteria consisted of patients over the age of 18 with a unilateral FP diagnosis of any etiology (Bell’s, congenital, vascular, cranioencephalic trauma, neoplastic or infectious). Exclusion criteria included any history of eyelid surgery and/or facial trauma. Patients or the public were not involved in the design, or conduct, or reporting, or dissemination plans of our research.

Disease staging was done using the House-Brackmann scale. DED evaluation consisted of the Ocular Surface Discomfort Index (OSDI) score, TMH, TBUT, meibography, and conjunctival hyperemia using an Oculus 5M Keratograph (®K5 M; Oculus GmbH, Wetzlar, Germany). Meibographies were later analyzed using ImageJ software (Research Services Branch, National Institute of Mental Health, Bethesda, Maryland, USA) to determine the percentage of glandular area loss. The sample calculation was done using a formula for estimating the proportion in an infinite population, p value of 0.0012 established 20 patients as an appropriate sample size.

## Results

In our sample, 11 patients (55%) were female, mean age of diagnosis was 57.80 ± 18.28 years (range of 22-80 years), and the evolution time of the disease was 67.05 ± 156.8 months (range of 3-720 months). Disease etiology was mostly Bell’s palsy, with 10 patients (50%) presenting as such, the rest of the etiologies are summarized in Table 1. In terms of laterality, the most affected side was the left hemiface in 13 patients (65%). Disease severity was predominantly grade IV in the House-Brackmann scale (8 patients, 40%) and the rest being reported in Graph 1.

**Graph S1.**
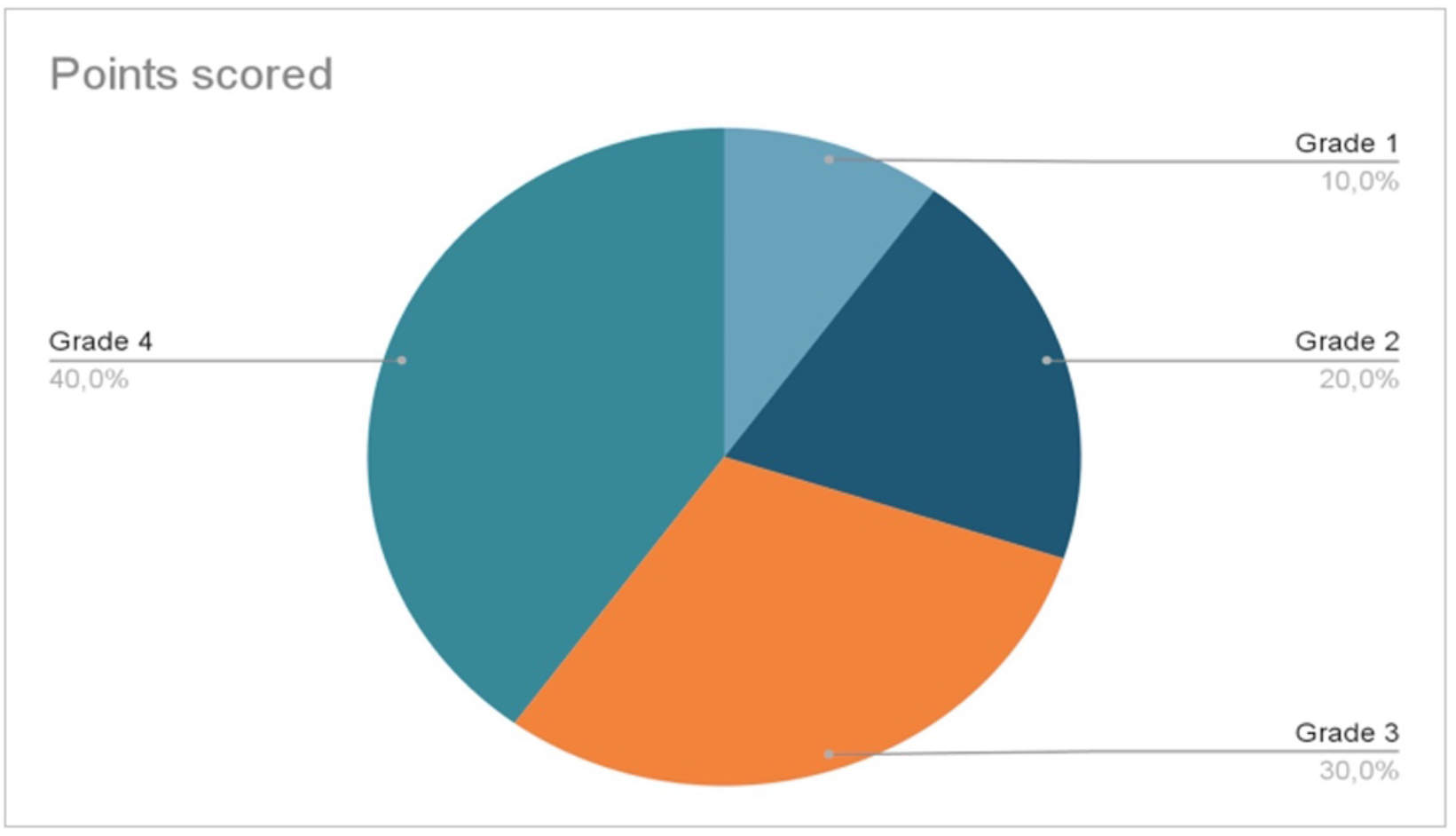
Disease severity.

**Table 1.** Facial Palsy etiologies reported in the study.

| Etiology | Number of patients |
| --- | --- |
| Bell's palsy | 10 (50%) |
| Iatrogenic | 5 (25%) |
| Cerebrovascular disease | 3 (15%) |
| Cranioencephalic trauma | 2 (10%) |

TF analysis using the 5M keratograph demonstrated a statistically significant difference (p=0.0199) in TMH when comparing sides, with an increased TMH in the affected side. Another finding was the difference in TBUT, where the paretic side presented a diminished TBUT compared to the healthy side, but it wasn’t statistically significant (p=0.2167). Conjunctival hyperemia was more evident in the affected side and was statistically significant (p= 0.0202) and OSDI questionnaire reported a mean score of 64.947 ± 19.79, which reflected a severe grade of dry eye symptomatology. Meibography analysis using ImageJ software yielded statistically significant results when comparing upper and lower Meibomian gland area percentage loss, especially in the lower eyelid. These findings are summarized in table 2.

**Table 2.** Meibography analysis using ImageJ.

| Area of loss | Affected side | Unaffected side | p value |
| --- | --- | --- | --- |
| % of Upper lid Meibomian glands | $29.55 \pm 13.31$ | $20.85 \pm 12.54$ | 0.0374 |
| % of Lower lid Meibomian glands | $44.44 \pm 16.9$ | $25.51 \pm 13.38$ | <0.001 |

## Discussion

Our study demonstrates the changes in TF dynamics and MGD in patients with unilateral FP and highlights the differences when comparing the affected vs unaffected sides. The epidemiology reported in our sample reflects the study by Tavares-Brito et al^34^, where 920 patients with FP were evaluated and most of them were females with Bell’s palsy as the most common etiology. ^34^ However, certain differences such as age of presentation and evolution time of the disease were observed, with both being greater in our study.

When assessing DED by JENVIS Dry Eye Report, we noticed a lack of research in the literature, but our findings reflect other results from studies by Wan et al^2^ and Altin Ekin et al^35^, where TBUT was decreased in the affected side when compared with the unaffected side. An interesting finding in our study was the increased TMH in the paretic side, since we could not find other studies reporting the same result. The differences in TMH between sides can be explained by the loss of muscular tone and increased laxity of the lower eyelid. Furthermore, since there is very little research on meibographies in patients with FP, this study contributes to the structural and qualitative analysis of these glands.

Another crucial factor that cannot be overlooked is the orbicularis oculi function, specifically Riolan’s muscle, in glandular dynamics. When blinking occurs, Riolan’s muscle contributes to Meibomian gland drainage, thus integrating the lipid component of the tear film.^8^ The importance of this layer lies in its ability to diminish the surface tension of the tear film, this prevents tear “spillage” over the palpebral margin. Also, the lipid component helps in preventing evaporation of the tear and improves lubrication and distribution over the eyeball. ^36,38-40^ Thus, when the paretic hemiface tries to blink, Riolan’s muscle cannot contract properly, and this leads to improper drainage of Meibomian glands. This study warrants further investigation on the specific degree of Riolan’s muscle disfunction.

The difference in glandular loss area when comparing the paretic vs non-paretic side was evident, as well as statistically significant. Although other factors contribute to Meibomian gland degeneration, such as hormones and environmental insults, this difference between healthy and affected sides may be explained by the FP itself. The disease mechanism consists of glandular obstruction and eventual atrophy due to incomplete blinking and Riolan’s muscle dysfunction.

^8,15^ Most of the patients in the study presented with a grade 4 in the House Brackman scale, which corresponds to incomplete blinking, and the effects of this are in accordance with previous findings ^41^. Although there are different MGD staging scales, evaluator experience plays an important role in detecting the percentage of glandular atrophy. In image 1, we present a female patient with right-sided FP; if the Meiboscore proposed by Arita et al^37^ is used, a grade of 1 and 0 can be established in the right and left upper eyelids, respectively. However, by using ImageJ software, we determined both as grade 1, thus demonstrating a clear difference between human and computer evaluation. This fact highlights potential meibomian gland misclassification without the aid of imaging software.

**Image 1.**
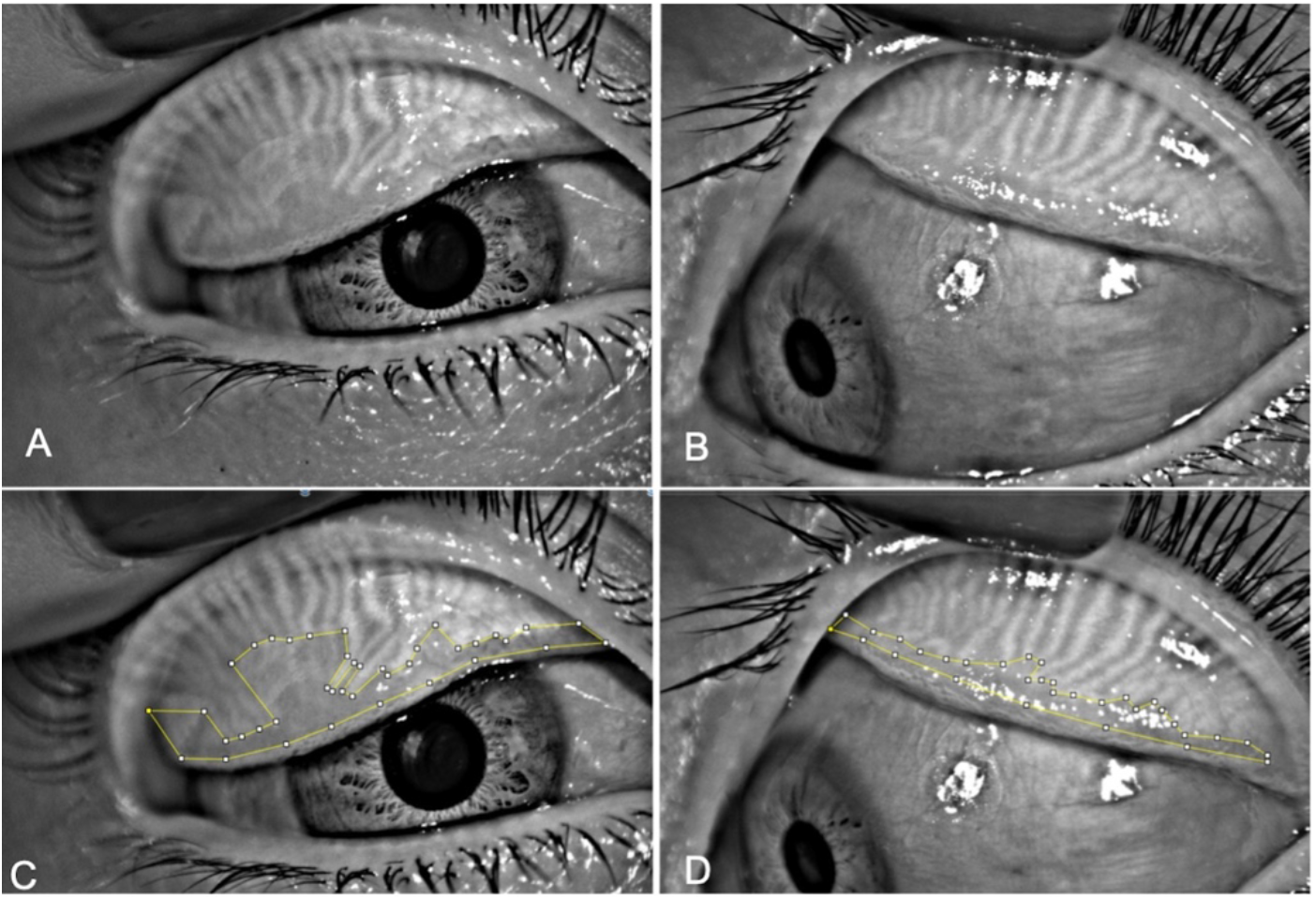
Upper eyelid Meibographies of a female patient in their 30s with right-sided facial palsy. Glandular loss area analysis made with ImageJ.

The degree at which the glands are affected also appears to be different depending on the vertical axis, with the lower lids being more affected when comparing them to their upper counterpart. In Image 2, a male patient presented a greater loss in the paretic side, but it was even more evident in the lower glands, where 50% loss was observed while the upper glands presented a loss of 28%. This may be explained by the increased laxity of the eyelid and the consequent exposure of the glands to environmental factors.

**Image 2.**
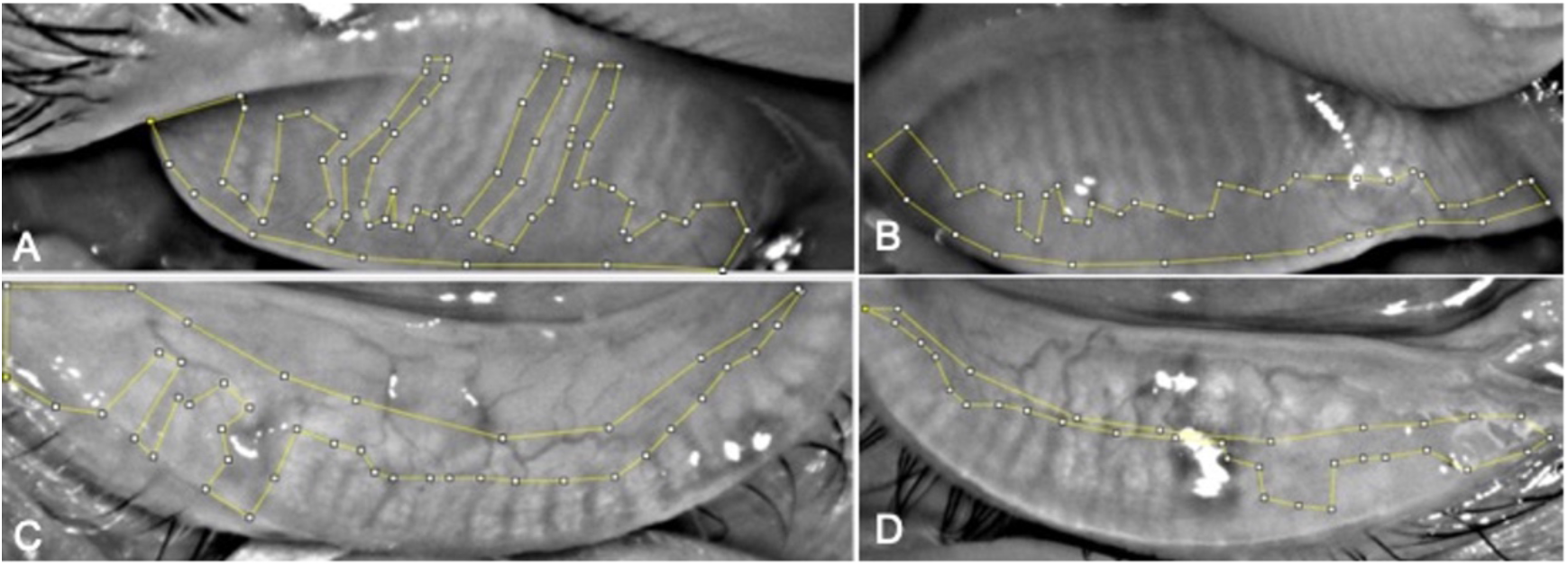
Lower eyelid meibographies of a male patient in their 70s with right-sided facial palsy. Glandular loss area analysis made with ImageJ.

In summary, a complete DED evaluation should be performed when examining patients with a FP diagnosis. It is crucial to evaluate the tear film in these patients, as well as the Meibomian glands, because of all the implications in quality of life that FP can entail. There is still much to be learned from the Meibomian glands in patients with FP, from their structural changes to their functional alterations. This study can expand its horizons even further, such as evaluating disease progression with different treatment modalities. A limitation in our study was the sample size due to the COVID-19 pandemic, but it stands to reason that an even greater sample could improve the statistical power of the study. Another limitation was the use of the House Brackman as the main grading of severity for facial palsy. Even though it is a mostly subjective tool, it was included because it is the most widely used grading scale for facial palsy, it is fast and relatively easy to do. The CADS score, an ophthalmology specific grading scale was tried to use, but many patients of our sample had already undergone eye surgery, which is an excluding factor in the scale. This grading scale offers several objective measures of periocular facial nerve function, and it will be surely utilized in the future.^42^

As previously stated, evaluator experience weighs heavily in determining MGD severity, and ImageJ analysis showed greater meiboscore and area of glandular loss when compared to the clinician’s evaluation. This is a valuable finding as it complements their diagnostic arsenal and may help guide in the decision of early diagnosis and treatment of MGD.

## Data Availability

All data produced in the present study are available upon reasonable request to the authors

## Acknowledgments

None

## Ethics approval statement

This study was approved by the local ethics committee (Comité de Ética Tecnológico de Monterrey) in February 2020 according to the Helsinki declaration under the registration number: P000302-CDLPPFU. Informed consent was obtained from all participants.

## Contributions

CGdF: Concept, protocol design, data collection, data analysis, writing paper. JIRA: patient evaluation, performing meibographies. DBF: data analysis, writing, review, and editing. AdCM: writing, review, and editing. F.V.V: Data collection. J.E.V.G: Concept, protocol design, writing, review, and editing. All authors have read and agreed to the final version of the manuscript.

